# SARS-CoV2 wild type and mutant specific humoral and T cell immunity is superior after vaccination than after natural infection

**DOI:** 10.1101/2021.09.07.21262725

**Authors:** Jennifer R. Richardson, Ralph Götz, Vanessa Mayr, Martin J. Lohse, Hans-Peter Holthoff, Martin Ungerer

**Author notes:** Correspondence to: Prof. Martin Ungerer, MD, or to Hans-Peter Holthoff, PhD, ISAR Bioscience, Semmelweisstrasse 5, D 82152 Planegg, Germany. shared senior authorship.

## Abstract

**Objective:** We investigated blood samples from fully SARS-CoV2-vaccinated subjects and from previously positive tested patients up to one year after infection with SARS-CoV2, and compared short- and long-term T cell and antibody responses, with a special focus on the recently emerged delta variant (B.1.617.2).

**Methods and Results:** In 23 vaccinated subjects, we documented high anti-SARS-CoV2 spike protein receptor binding domain (RBD) antibody titers. Average virus neutralization by antibodies, assessed as inhibition of ACE2 binding to RBD, was 2.2-fold reduced for delta mutant vs. wild type (wt) RBD. The mean specific antibody titers were lower one year after natural infection than after vaccination; ACE2 binding to delta mutant vs. wt RBD was 1.65-fold reduced. In an additional group, omicron RBD binding was reduced compared to delta.

Specific CD4+ T cell responses were measured after stimulation with peptides pools from wt, alpha, beta, gamma, or delta variant SARS-CoV2 spike proteins by flow cytometric intracellular cytokine staining. There was no significant difference in cytokine production of IFN-γ, TNF-α, or IL-2 between vaccinated subjects. T cell responses to wt or mutant SARS-CoV2 spike were significantly weaker after natural occurring infections compared to those in vaccinated individuals.

**Conclusion:** Antibody neutralisation of the delta mutant was reduced compared to wt, as assessed in a novel inhibition assay with a finger prick blood drop. Strong CD4 T cell responses were present against wt and mutant SARS-CoV2 variants, including the delta (B.1.617.2) strain, in fully vaccinated individuals, whereas they were partly weaker 1 year after natural infection. Hence, immune responses after vaccination are stronger compared to those after naturally occurring infection, pointing out the need of the vaccine to overcome the pandemic.

## Introduction

Since December 2019, the SARS-CoV-2 pandemic has caused global health problems, leading to more than 4 million deaths (Johns Hopkins University database) and therefore demanding rapid development of vaccines for protection against the virus. Vaccine development has included mRNA, viral vectors, recombinant proteins and inactivated virus (1), leading to discussions about the efficiency of the various approaches in terms of humoral and cellular immune responses against the virus. Several vaccines have been investigated in large clinical trials and have demonstrated safety and efficacy (2-4): among them, BNT162b2 (Pfizer-BioNTech; mRNA), mRNA-1273 (Moderna; mRNA), and ChAdOx1 nCoV-19 (AZD1222) (Oxford-AstraZeneca; chimpanzee adenoviral vectored) have been approved globally and have been most frequently used in Europe. All of these vaccines have been designed to raise antibodies and T lymphocyte responses to the spike (S) protein. All include S sequences derived from the first reported sequence from Wuhan in January 2020 (5).

Similar to other RNA viruses, SARS-CoV-2 is subject to progressive mutational changes. The pandemic spread leads to a huge extent of viral replication, increasing the odds that adaptive mutations will occur, which could lead to selective advantages, e.g. enhanced binding to human cells or immune escape from neutralizing antibodies (6). The S protein is a type-1 transmembrane glycoprotein, which may assemble into trimers (7). It is composed of two parts: the S1 domain bears the receptor-binding domain (RBD) and mediates cell binding via the angiotensin-converting enzyme-2 (ACE2), while the S2 domain completes membrane fusion, allowing the viral RNA to access the host cell cytoplasm to initiate viral replication.

The ACE2-RBD interaction is mediated by a small 25 amino acid patch, which becomes accessible when the RBD moves into an upper direction (8; 9). Mutations in this region are most concerning:

The alpha (B.1.1.7), beta (B.1.351), gamma (P.1) and delta (B.1.617.2) variants have been classified as variants of concern and have by far superseded the wild type (wt) strain. All of these strains have the potential to modulate ACE2-RBD binding affinity, potentially leading to an increased transmissibility. In addition, the variants’ mutated amino acid residues can also modulate neutralization of SARS-CoV2 by naturally or vaccine induced antibody responses. Neutralisation has been assessed by using live (pseudo)virus which infects living cells, and measure inhibition by adding test serum samples. This method can be successfully replaced by an assay which determines inhibition of RBD-ACE2 binding (10).

In this study, we therefore compared the immune responses of individuals less than 2 months or 1 year after a naturally occurring infection with SARS-CoV2 with those of individuals who have been completely vaccinated (at least two weeks after the last dose) with either a mRNA-based (BNT162b2, Comirnaty (Pfizer-BioNTech), mRNA-1273 (Moderna)) or a vector-based (ChAdOx1 nCoV-19 - Vaxzevria, AZD1222, Oxford-AstraZeneca) vaccine. We investigated anti-S and anti-N protein antibody titers and performed inhibition assays of ACE2 binding to wildtype vs. delta mutant RBD proteins (as a correlation of neutralizing potency). Further, S protein-specific T cell responses, determined by cytokine production after stimulation with wt and mutant S protein-derived peptide pools were measured. The study was intended to provide further insight into immune responses after naturally occurring infections versus vaccination regarding the most concerning mutant virus variants with a special focus on the delta mutant.

## Results

### Protein production and purification

SARS-CoV2 mutant spike RBD and nucleocapsid wild type and mutant proteins were expressed in mammalian cells and purified to > 95% purity (by mass spec and Coomassie staining) to be used in the assays. Figures 1A + B show representative examples for the purity after the affinity column and after subsequent dialysis, respectively.

**Figure 1:**
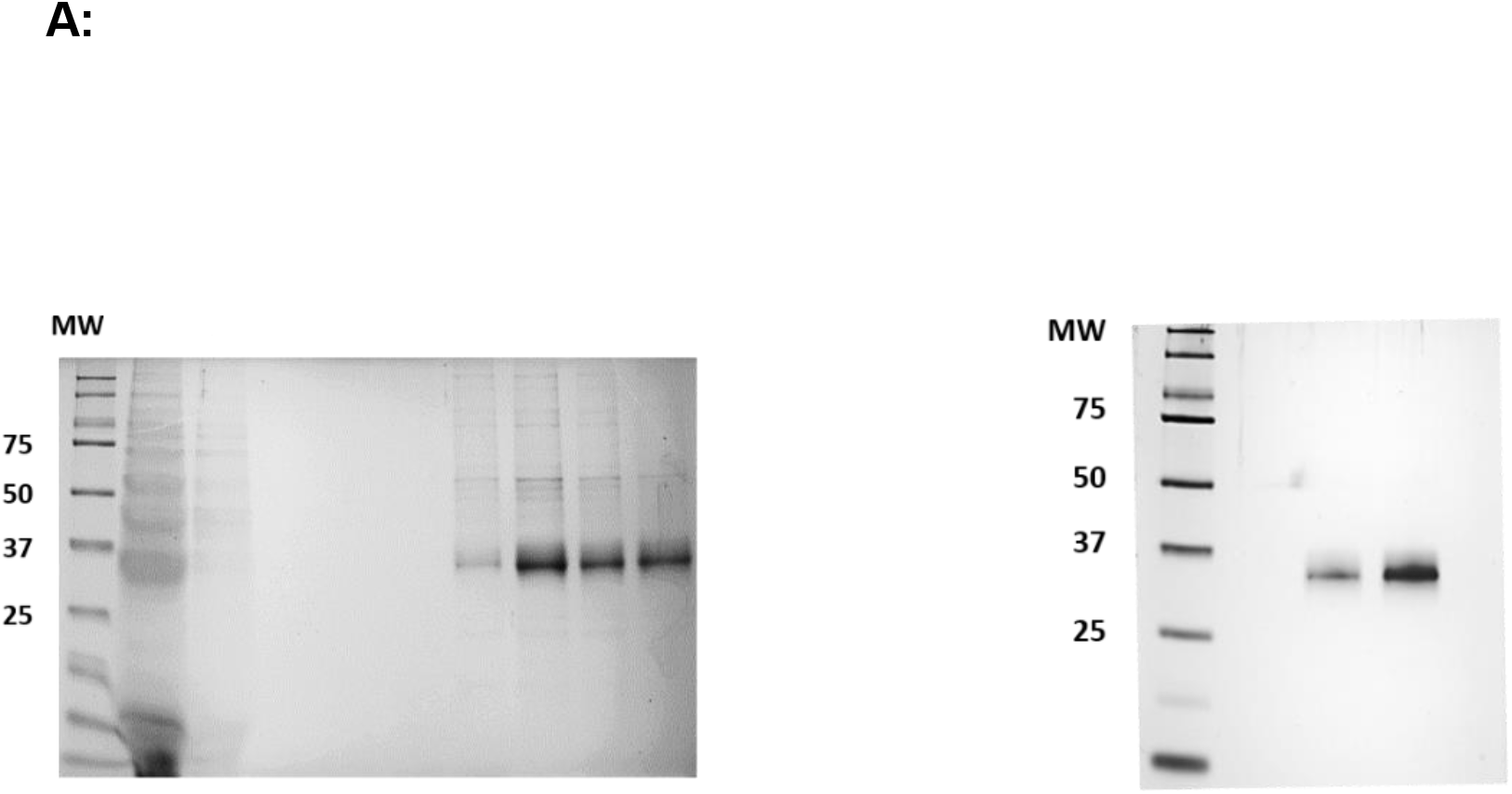
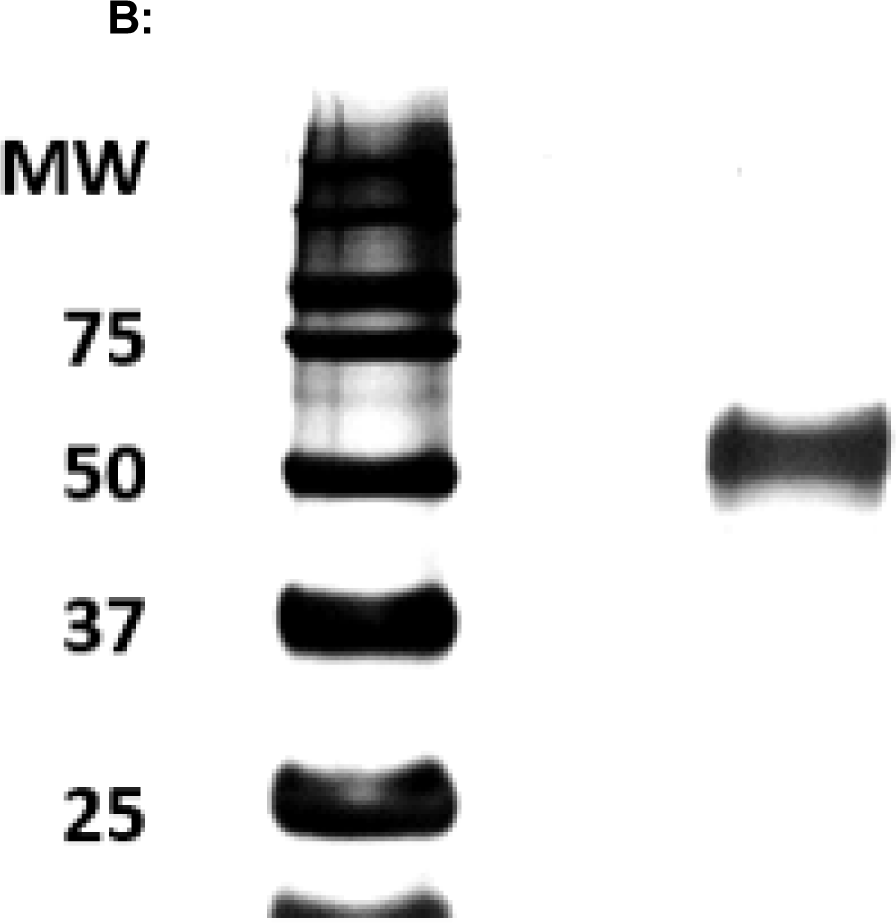
**A:** Representative Coomassie gel electrophoresis (left panel) and silver stain (right panel) of SARS-CoV2 wild type spike RBD protein (A) and of nucleocapsid protein (B).

### Anti-RBD spike and anti-nucleocapsid protein antibody titers

All vaccinees showed clear antibody responses against wildtype SARS-CoV2 RBD regardless of the type of vaccine used – as shown in Figure 2A. We did not detect any anti-nucleocapsid protein antibodies in any subject vaccinated with BNT162b2, except in one, who had suffered from naturally occurring infection before vaccination, with an anti-N titer of 28 arbitrary units (AU). In addition, one subject vaccinated with mRNA-1273 (Moderna) showed anti-N antibodies with a titer of 29 AU.

**Figure 2:**
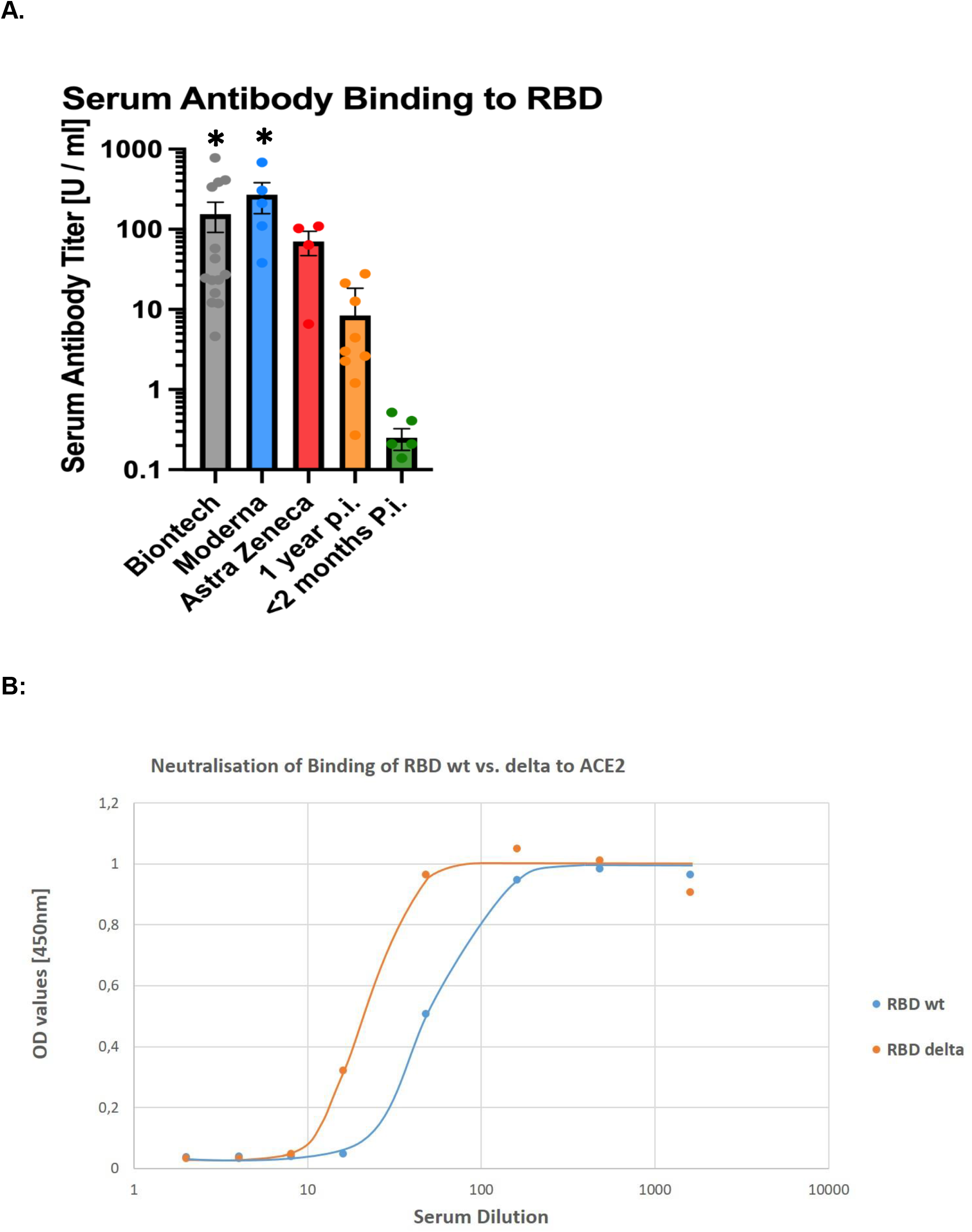
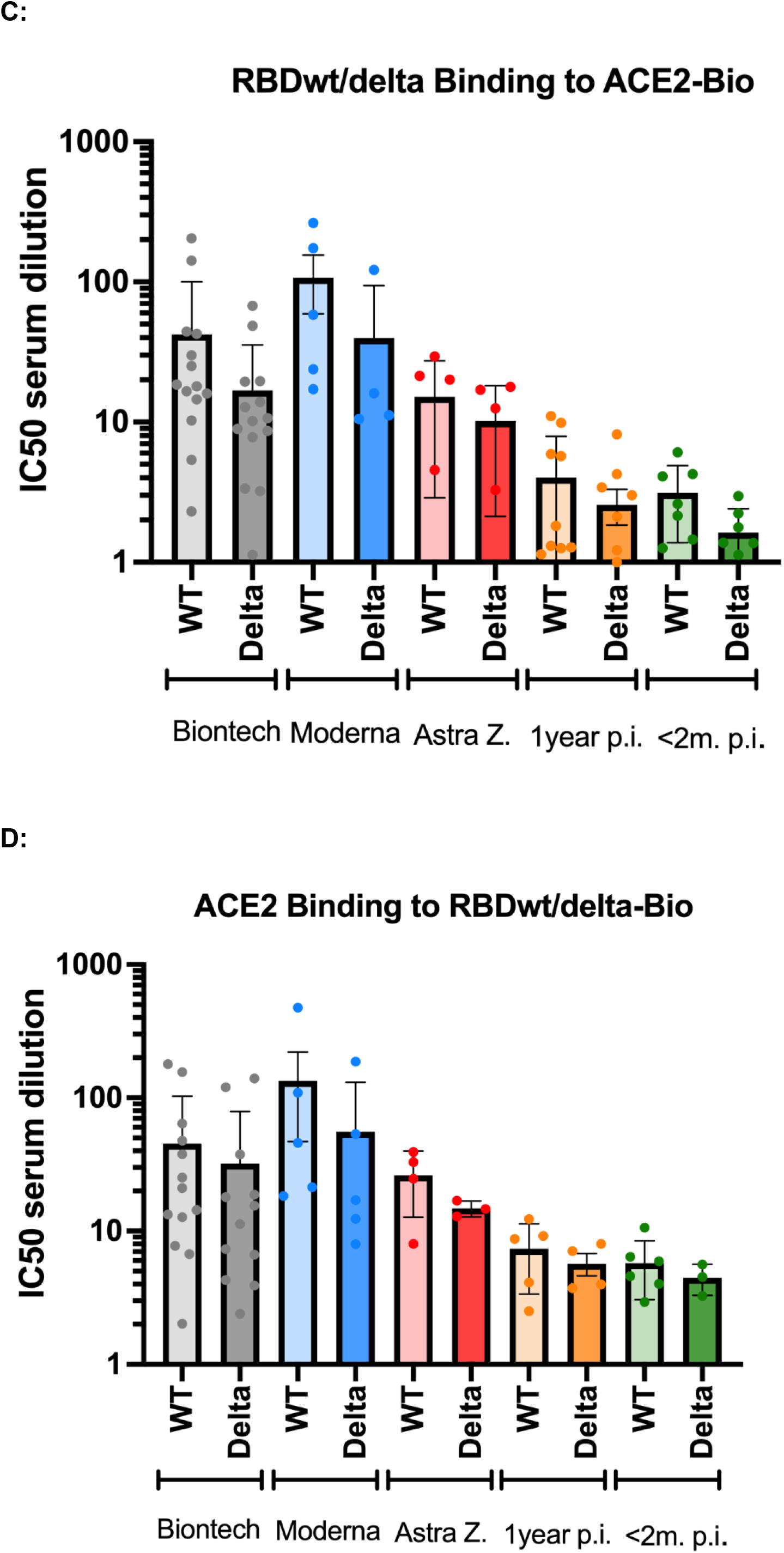
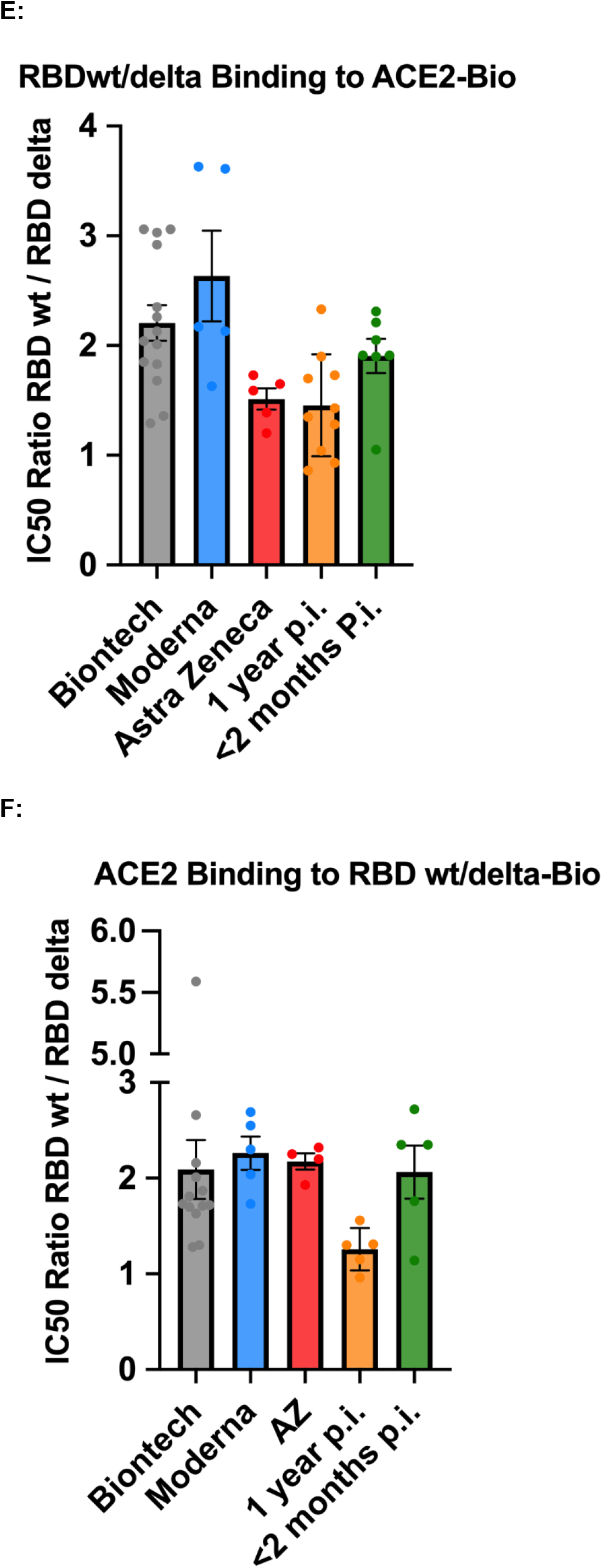
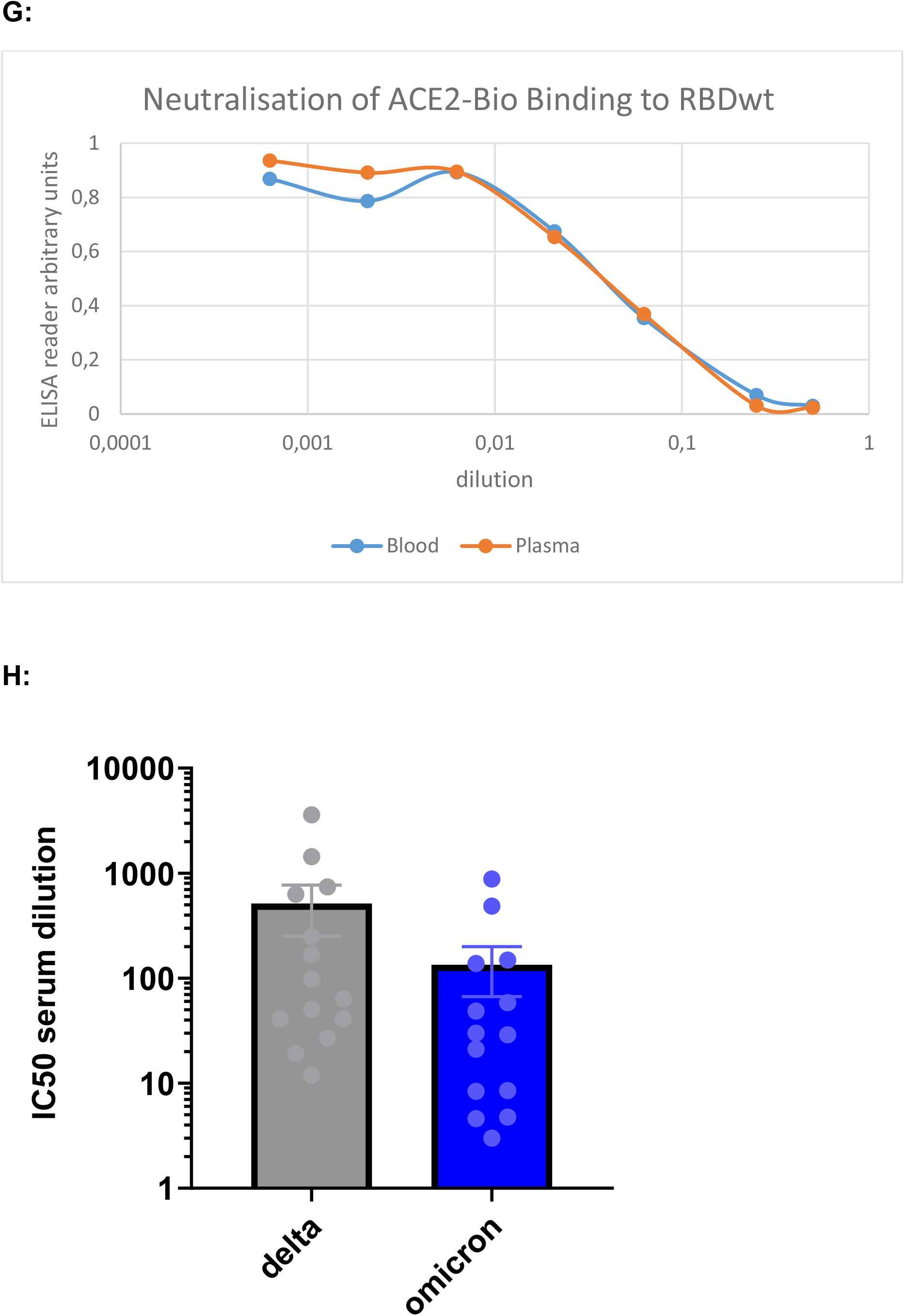
Serum Antibodies and Neutralisation. The bar graphs show results of n=13 independent individuals after vaccination with BioNtech (grey), n=5 Moderna (blue), n=5 AstraZeneca (red), and n=10 independent individuals one year (orange), and n=7 subjects < 2 months (green) after natural infection with standard errors of the mean (SEM). Significance was tested by one-way ANOVA (A) or by Kruskal-Wallis test for non-parametric values (B-E). **p < 0.05 vs. naturally infected subjects.**A:** Anti RBD antibody titres; **B:** Representative curve of a single experiment to determine IC50-values for the inhibition of biotinylated ACE2 binding to immobilized wt vs. delta mutant RBD. **C:** Absolute IC50-values calculated for the inhibition of biotinylated ACE2 binding to immobilized wt vs. delta mutant RBD. **D:** Absolute IC50-values calculated for the inhibition of biotinylated wt vs. delta mutant RBD binding to immobilized ACE2. **E:** Ratios between IC50-values calculated for inhibition of ACE2 binding to immobilized wt vs. delta mutant RBD. **F:** Ratios between IC50-values calculated for inhibition of wt vs. delta mutant RBD binding to immobilized ACE2. **G:** Read-out of a representative experiment showing the comparison between inhibition of biotinylated ACE2 binding to immobilized RBD with serum taken by venous sampling vs a drop of full blood. **H:** In additional, independent group of 14 subjects who had been fully vaccinated with BNT 162b2, we also investigated ratios between IC50 values calculated for inhibition of ACE2 binding to immobilized omicron vs. delta mutant RBD.

Further, we compared short-term and long-term humoral immune responses in patients’ blood samples up to one year after infection with SARS-CoV2 (Figure 2A). We found specific anti-SARS-CoV2 RBD spike protein antibody titers in 90% of the investigated individuals, as assessed by comparing these titers with non-specific protein binding on the plates. Titers in individuals who had received mRNA vaccines were significantly higher than in subjects after natural infection, regardless of the time interval since infection.

In addition, we detected anti-nucleocapsid protein antibodies in all patient blood samples in these 2 groups: anti-N titer amounted to 66±21 AU one year and to 21±9 AU 2 months after natural infection.

### RBD – ACE2 inhibition as a correlate of neutralizing antibodies

The inhibition of either wildtype SARS-CoV2 RBD or delta mutant RBD binding to ACE2 was investigated as a correlate for antibody neutralizing potency. A representative result curve from a single experiment is shown in Figure 2B. We found significantly reduced mean IC50-values for the delta mutant – see Figures 2C-F. Figure 2C shows absolute IC50-values for the inhibition of ACE2 binding to immobilized wt vs. delta mutant RBD.

Figure 2D shows absolute IC50-values calculated for the inhibition of wt vs. delta mutant RBD binding to immobilized ACE2. The ratios between IC50-values calculated for inhibition of ACE2 binding to immobilized wt vs. delta mutant RBD are shown in Figure 2E. Figure 2F shows ratios between IC50-values calculated for inhibition of wt vs. delta mutant RBD binding to immobilized ACE2. Both, vaccinees and naturally infected subjects showed significantly (p < 0.001 by sign testing) reduced mean IC50-values for the delta mutant, with an average 2.2-fold reduction in vaccinees and an 1.65-fold average reduction in the naturally infected group. Figure 2G shows that results using a drop of finger-stick full blood (about 30 µl) were well comparable to those using sera obtained by venous puncture.

In summary, inhibition assays yielded similar results, regardless whether RBD or ACE2 was coated onto plates, and whether RBD or ACE was biotinylated and added in solution – and that the assay can be performed with just a drop of finger-stick blood.

As another variant of concern, omicron, emerged during the review process of this manuscript in December 2021, we include additional measurements of the relative IC50 values of omicron versus delta in an additional, independent group of 14 subjects who had been fully vaccinated with BNT 162b2. The results (Figure 2H) show decreased values for omicron, as expected, thus providing another proof of concept for the method.

### Antigen-specific CD4+ T cell responses against the spike protein

In a first experimental set-up, CD4+ T cell responses of vaccinated as well as naturally infected individuals were investigated using a wild type SARS-CoV2 S protein peptide pool derived from the whole amino acid sequence of wt protein which was available from the company Miltenyi at the time of this first study. CD4+ T cell responses were measured by intracellular cytokine staining.

Frequencies of interferon-γ (IFN-γ)-positive, tumor necrosis factor-α (TNF-α)-positive, or interleukin-2 (IL-2)-positive cells are shown in figure 3. Significant T cell responses were observed both after vaccination and after natural infection compared to the negative controls (individuals who had not been vaccinated or infected). Using the Miltenyi peptide pool, we observed higher mean T cell responses in individuals who had been vaccinated with the Moderna mRNA formulation in this cohort.

**Figure 3:**
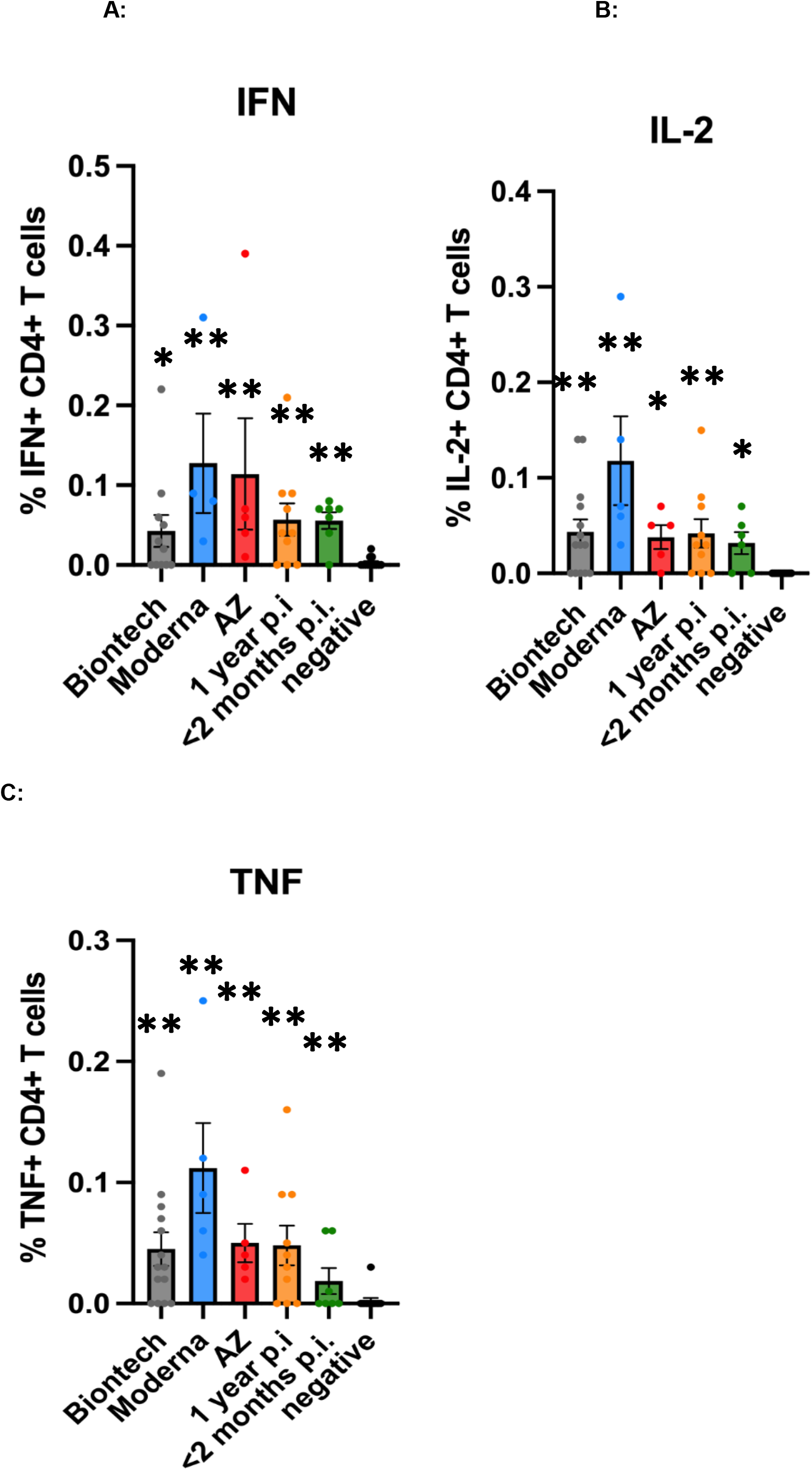
Antigen-specific CD4+ T cell responses to the Miltenyi peptide S protein pool. Antigen-specific CD4+ T cell responses elicited by the Miltenyi wt S protein peptide pool after vaccination with BioNtech (grey), Moderna (blue) or AstraZeneca (red), or one year (orange) or < 2 months (dark green) after naturally occurring infection with SARS-CoV2. In comparison, samples of healthy control subjects, who tested negative for SARS-CoV2-antibodies, are shown on the right of each panel. The bar graphs show frequencies of (A) interferon-γ (IFN-γ), (B) interleukin-2 (IL-2)- and (C) tumor necrosis factor-α (TNF-α)-positive CD4+ T cells. The Figure shows results of n=13 independent individuals after vaccination with BioNtech, n=5 Moderna, n=5 AstraZeneca, and n=10 independent individuals one year and n=7 subjects < 2 months after natural infection with standard errors of the mean (SEM). For comparison, 13 negative healthy controls were included. Significance levels were tested by Kruskal-Wallis test for non-parametric values. *p < 0.05, **p < 0.01 vs. healthy controls

A follow-up experimental set-up then compared the CD4+ T cell responses against peptide pools whose sequences comprised the whole amino acid sequence of wild type and alpha, beta, gamma and delta spike protein mutants, which were obtained from JPT peptides (Figures 4 – 6). Frequencies of interferon (IFN)-gamma (IFN-γ)+ CD4+ cells (Figure 4), of interleukin (IL-2+ CD4+ cells (Figure 5) and of tumour necrosis factor-alpha TNF-α) (Figure 6) for all subjects were determined by intracellular cytokine staining. No differences in the frequencies of cytokine-positive CD4+ T cells were observed between the different S peptide pools in one group. The mRNA vaccines, however, yielded higher frequencies of cytokine positive CD4+ T cells compared to the Astra Zeneca vaccine and the naturally occurred infections.

**Figure 4:**
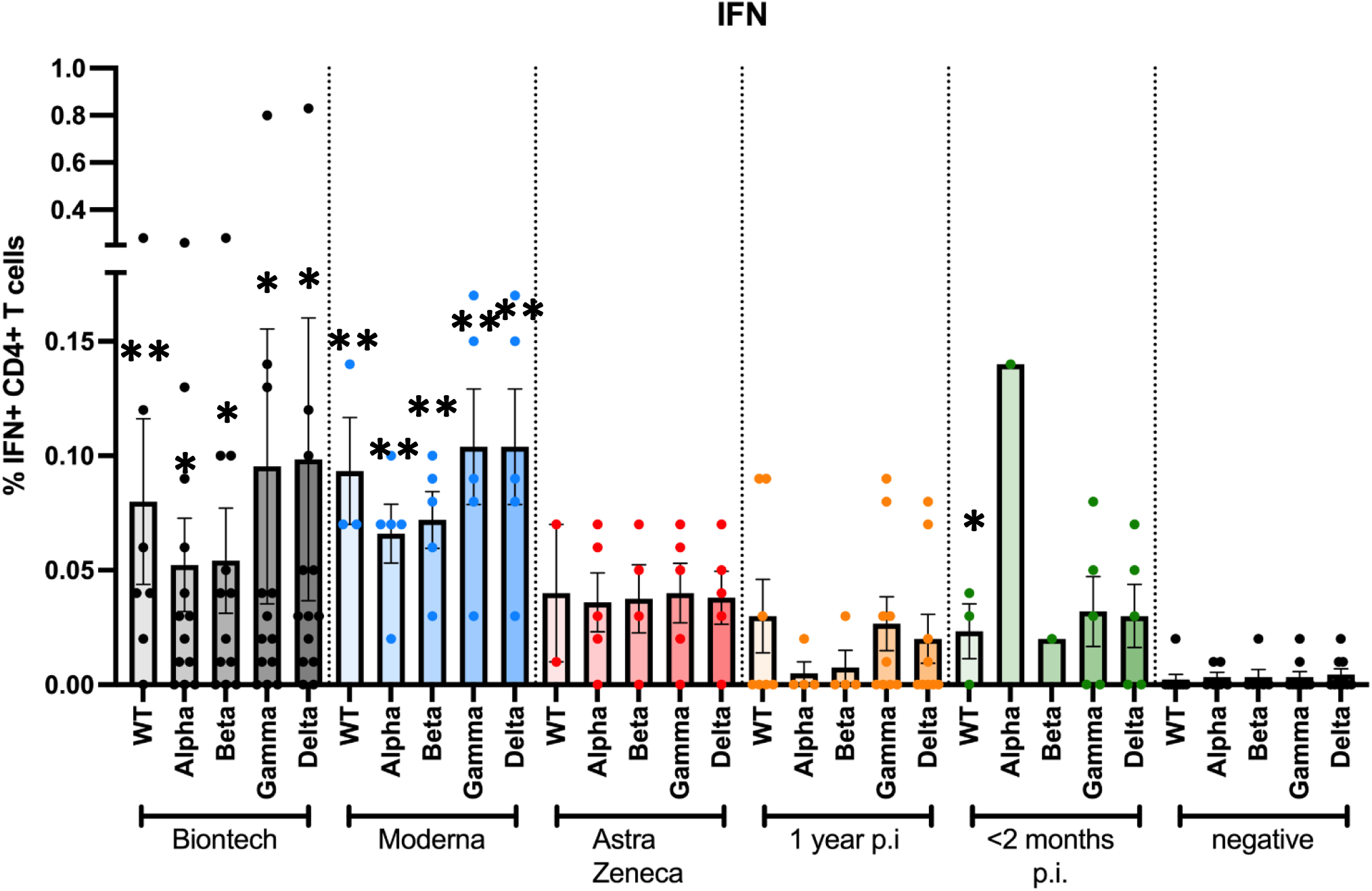
Antigen-specific IFN-γ CD4+ T cell responses to JPT peptide S protein wt and mutant pools. Antigen-specific CD4+ T cell responses elicited by the JPT peptide S protein pools after vaccination with BioNtech (grey), Moderna (blue) or AstraZeneca (red), or one year (orange) or < 2 months (dark green) after naturally occurring infection with SARS-CoV2, or negative controls. The bar graphs show frequencies of IFNγ+CD4+ T cells for wild type, and for alpha (B.1.1.7), beta (B.1.351), gamma (P.1) and delta (B.1.617.2) mutants. The Figure shows results of n=13 independent individuals after vaccination with BioNtech, n=5 Moderna, n=5 AstraZeneca, and n=10 independent individuals one year and n=7 subjects < 2 months after natural infection, and n=9 negative controls, with standard errors of the mean (SEM). Significance levels were tested with Kruskal-Wallis test for non-parametric samples. *p < 0.05 vs. healthy controls; **p < 0.01 vs. healthy controls and p < 0.05 vs 1 year after infection

**Figure 5:**
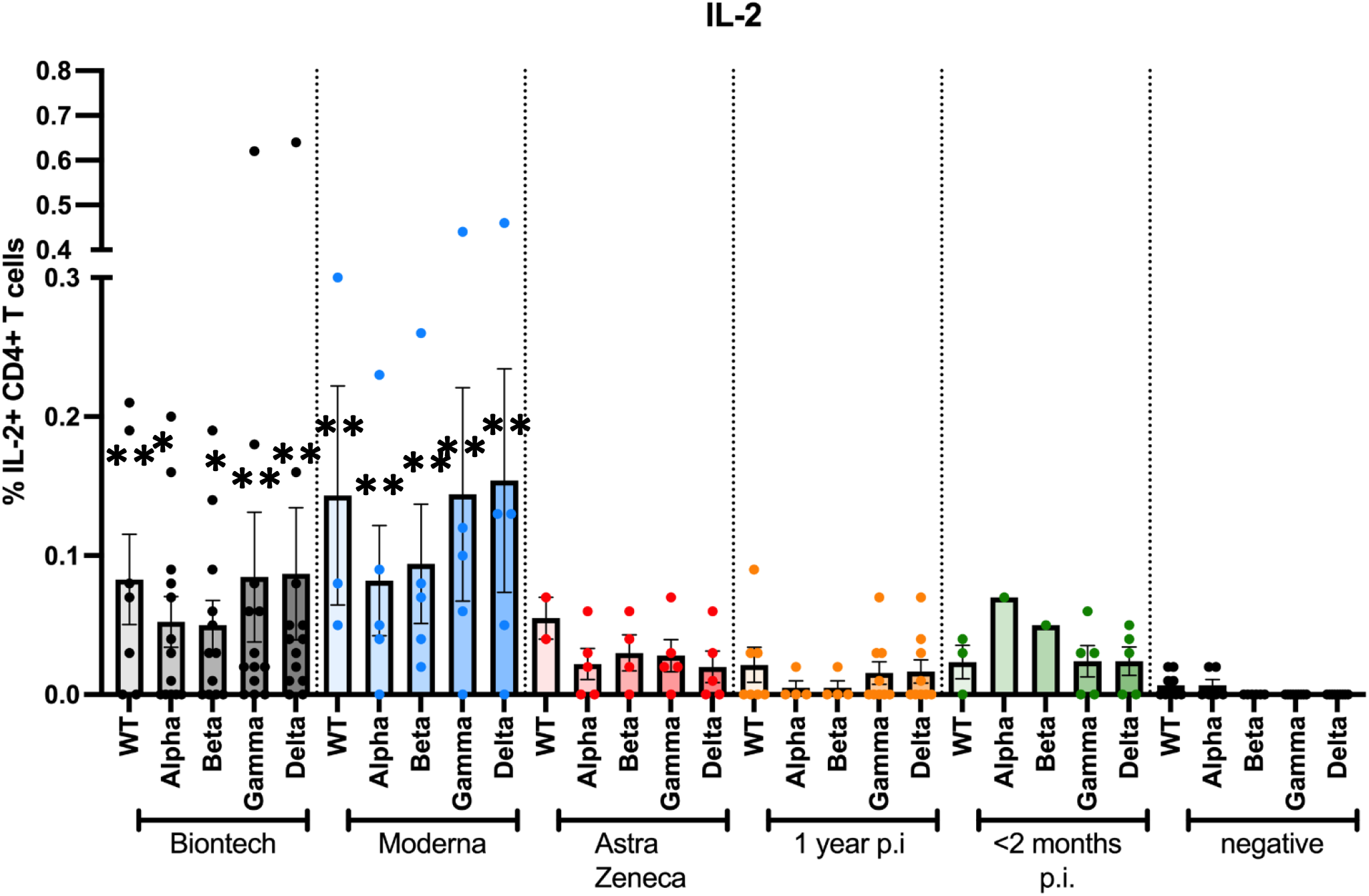
Antigen-specific IL2+ CD4+ T cell responses to JPT peptide S protein wt and mutant pools. Antigen-specific CD4+ T cell responses elicited by the JPT peptide S protein pools after vaccination with BioNtech (grey), Moderna (blue) or AstraZeneca (red), or one year (orange) or < 2 months (dark green) after naturally occurring infection with SARS-CoV2, or negative controls. The bar graphs show frequencies of interleukin-2 (IL-2)-positive CD4+ T cells for wild type, and for alpha (B.1.1.7), beta (B.1.351), gamma (P.1) and delta (B.1.617.2) mutants. The Figure shows results of n=13 independent individuals after vaccination with BioNtech, n=5 Moderna, n=5 AstraZeneca, and n=10 independent individuals one year and n=7 < 2 months after natural infection with standard errors of the mean (SEM). Samples of non infected individuals tested negative (not shown in the Figure to enhance legibility). Significance levels were tested by Kruskal-Wallis test for non-parametric values. **p < 0.01 vs. healthy controls, and p < 0.05 vs. 1 year after infection

**Figure 6:**
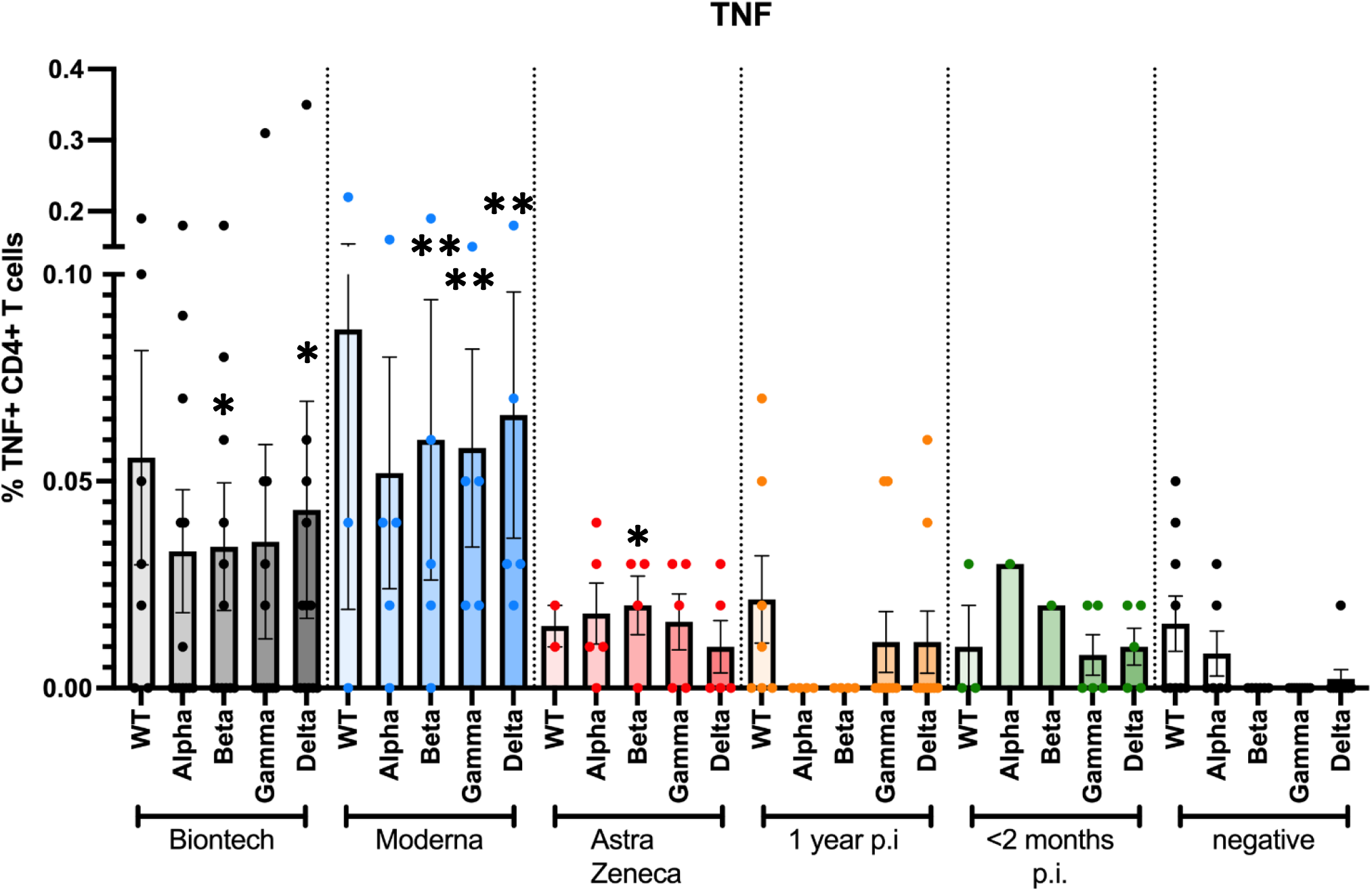
Antigen-specific TNF-α+ CD4+ T cell responses to JPT peptide S protein wt and mutant pools. Antigen-specific CD4+ T cell responses elicited by the JPT peptide S protein pools after vaccination with BioNtech (grey), Moderna (blue) or AstraZeneca (red), or one year (orange) or < 2 months (dark green) after naturally occurring infection with SARS-CoV2, or negative controls. The bar graphs show frequencies of TNF-α+CD4+ T cells for wild type, and for alpha (B.1.1.7), beta (B.1.351), gamma (P.1) and delta (B.1.617.2) mutants. The Figure shows results of n=13 independent individuals after vaccination with BioNtech, n=5 Moderna, n=5 AstraZeneca, and n=10 independent individuals one year and n=7 < 2 months after natural infection with standard errors of the mean (SEM). Samples of non infected individuals tested negative (not shown in the Figure to enhance legibility). At least 200,000 events were counted for each individual condition. Significance levels were tested by Kruskal-Wallis test for non-parametric values. *p < 0.05 vs. healthy controls, **p < 0.01 vs. healthy controls and also p < 0.05 vs. 1 year after infection

Specific antibody reactivities and CD4+ T cell responses against SARS-CoV2 mutants alpha, beta, gamma and delta were comparable to wt in all individuals, with no evidence for reduced responsiveness to any of the mutants (Figures 2 and 4 - 6). In comparison, samples of non-infected healthy individuals did not show specific CD4+ T cell responses, except for wt- and alpha-induced TNF-α responses. Significantly weaker CD4+ T cell responses were observed 1 year after naturally occurring infection than after vaccination: especially, alpha-, beta-, gamma and delta mutant S protein peptide-induced IFN-γ responses, but also alpha-, beta- and delta mutant-induced IL-2 responses and beta- and delta-induced TNF-α responses were significantly lower in subjects who had been infected 1 year before.

### Antigen-specific CD8+ T cell responses against the spike protein

CD8+ T cell responses of vaccinated as well as naturally infected individuals were also investigated using a wild type SARS-CoV2 S protein peptide pool derived from the whole amino acid sequence of wt protein which was available from the company Miltenyi at the time of this first study. CD8+ T cell responses were measured by intracellular cytokine staining (Figure 7A).

**Figure 7:**
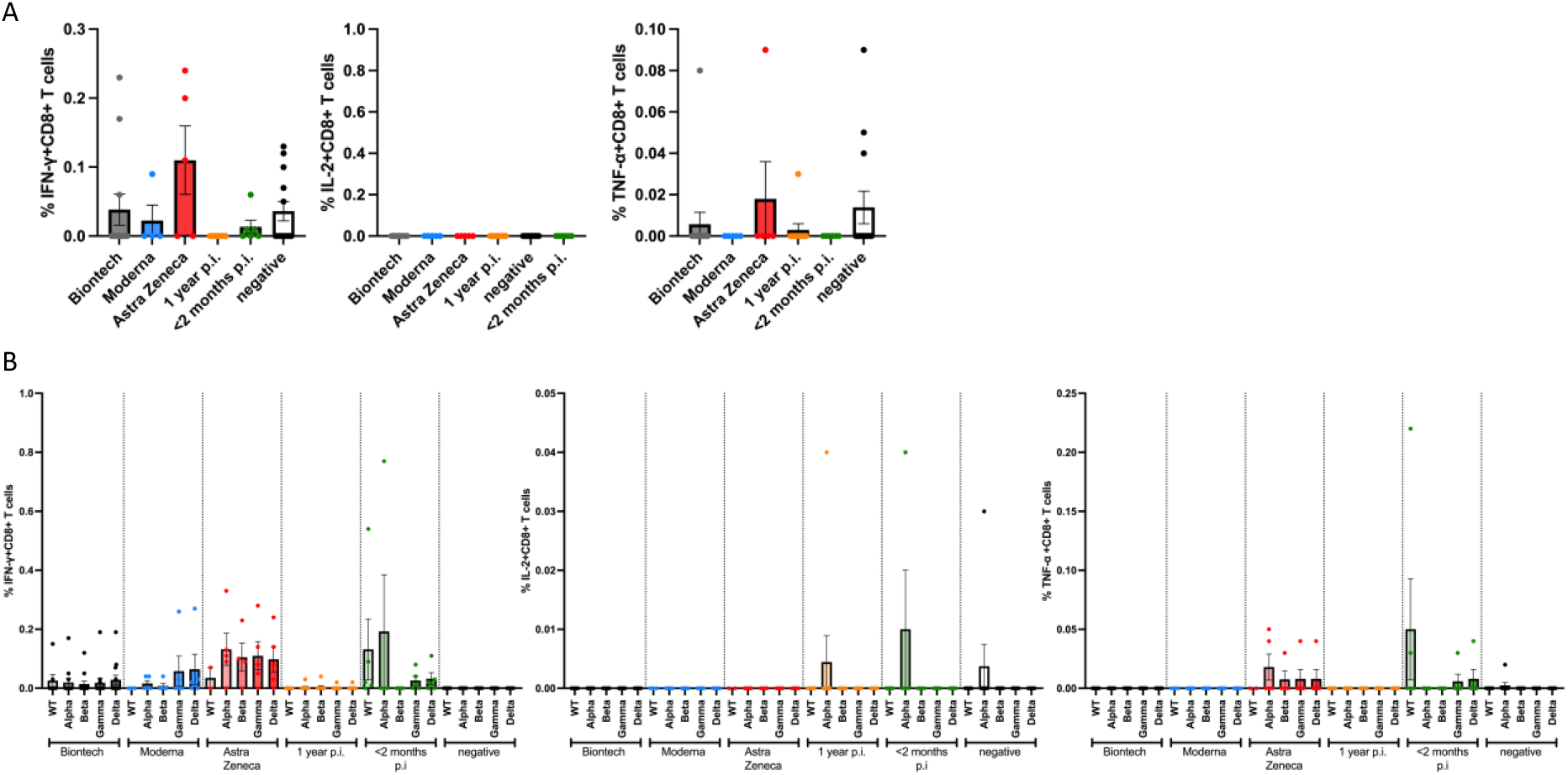
Antigen-specific CD8+ T cell responses to JPT and Miltenyi peptide S protein wt and mutant pools. Antigen-specific CD8+ T cell responses elicited by the JPT peptide S protein pools after vaccination with BioNtech (grey), Moderna (blue) or AstraZeneca (red), or one year (orange) or < 2 months (dark green) after naturally occurring infection with SARS-CoV2, or negative controls. The bar graphs show frequencies of TNF-α+, IL-2+ or IFNγ+ CD8+ T cells for wild type, and for alpha (B.1.1.7), beta (B.1.351), gamma (P.1) and delta (B.1.617.2) mutants. At least 200,000 events were counted for each individual condition.

A follow-up experimental set-up then compared the CD8+ T cell responses against peptide pools whose sequences comprised the whole amino acid sequence of wild type and alpha, beta, gamma and delta spike protein mutants, which were obtained from JPT peptides (Figure 7B). IFN-γ, IL-2 and TNF-α CD8+ T cells responses were analyzed for all the used peptide pools.

The frequencies were too low to be analyzed. These results corroborate those of other recent studies on CD8+ effector T cell responses after stimulation with pools of 15-meric epitope peptides, which we used in our study (12): These peptide samples are intended to measure CD4+ cells, but do not work appropriately for CD8+ cells.

The correlation between individual specific CD4+ T cell responses and anti-RBD antibodies was low, probably because B and T cell interaction is not linear, as determined by Pearson’s correlation testing or Spearman’s rank correlation, where appropriate.

## Discussion

All individuals who were vaccinated against SARS-CoV2 showed strong humoral immune reactions in terms of serum antibody binding to wild type (wt) and mutant virus proteins. A strong cellular CD4+ T cell immune reactivity against wt and mutant variant 15-meric peptide pools, including significant CD4+ T cell responses against the recently emerged gamma (P.1) and delta (B.1.617.2) variants was also detected. A specific CD8+ T cell response was, however, not detected, which might be due to the use of a 15-meric peptide pool, which is not sufficient to stimulate CD8+ T cells. Most responses were higher than those of patients up to one year after naturally occurring infection. Especially the results on the delta variant show that a combination of humoral response analysis based on rapid mammalian cell protein expression and adaptive T cell assays allows for quick investigation of immune responses to virus variants within 3 months after their emergence – which may be done even quicker for future variants of concern, as shown in our recent interim study on omicron RBD-ACE2 interaction.

Previous studies investigated antibody responses against SARS-CoV2 after COVID-19, and found that antibody titers decrease over time, which was assumed to correlate with decreasing immunity (12,13). Several recent studies investigated the long-term immune response after naturally occurring SARS-CoV2 infection or vaccination more broadly, including CD4+ helper and CD8+ effector T cells, memory B cells and others (14-16). Following up on these recent studies, we investigated another cohort in the Munich area, Bavaria, Germany. Specifically, we measured antibody and T cell responses against the S protein of recent variants of concern.

Our results show reduced inhibition of the delta mutant RBD – ACE2 interaction compared to wt RBD – ACE2 interaction by neutralizing antibodies. To our knowledge, our study systematically compared for the first time inhibition assays which used labelled wt vs. mutant RBD and coated ACE2 on plates with those in which ACE2 was labelled and added in solution to wt vs. mutant RBD-coated plates. The results were comparable for the specific assay conditions which we established – this novel approach enables wide-scale multiplex-compliant assays. Both the direct interaction of anti-SARS-CoV2 serum antibodies of vaccinated or naturally infected subjects with RBD, and the inhibition exerted by these antibodies on wt versus delta mutant RBD – ACE2 binding were measured by ELISA. The results using a drop of finger-stick full blood (about 30 µl) were well comparable to those using sera obtained by venous puncture, indicating that the assay can be up-scaled to a rapid and less invasive point-of-care ELISA.

Our findings corroborate other recent studies which investigated delta mutant RBD by assays using live (pseudo)virus (17-21), and resulted in comparable ratios of delta mutant versus wt binding. However, these assays are time-consuming and require specialized laboratory know-how. Although assessment of neutralization using in vitro binding assays may not reliably predict the potency of immune protection afforded by natural infection or immunization by real SARS-CoV-2, it can be carried out in non-specialized laboratories, and recent results using authentic virus largely substantiate the comparability (22). In over 90% of vaccinated individuals, neutralizing titers (NT50) remained well above the level associated with immune protection in recent vaccine trials (22,23). However, in many individuals post COVID-19 and recipients of a single dose of vaccine – neutralization capacity dropped (22). Reduced cross neutralization was observed against many virus variants and may impact on vaccine efficacy.

Generally, reduced anti-virus antibody neutralization as observed *in vitro* does not directly prove vaccine failure (24-26), because other immune mechanisms, especially T cell responses, also contribute significantly to virus inactivation and elimination. They often follow a different time course. For example, infection with influenza virus results in reduced disease upon subsequent infection with other strains, in both human and animal challenge studies, implying the importance of cellular immune mechanisms on protection elicited by vaccination (25).

Therefore, we also investigated T cell responses using peptide pools which were derived from the whole S sequence. This approach allowed for a broader investigation of anti-S protein immune responses. Generally, T cell responses against SARS-CoV2 are known to target a wide range of regions in the spike protein (27, 28). We found that significant immune responses were elicited by all investigated vaccines including mRNA (Moderna and BioNtech), and the vector-based vaccination with Vaxzevria (AstraZeneca). Our results show strong and comparable specific T cell responses to alpha (B1.1.7), beta (B1.351), gamma (P.1) and delta (B.1.617.2) spike protein variants in vaccinated subjects after two doses. Surprisingly, responses after vaccination with BioNtech were lower when tested with the initial Miltenyi than with the JPT peptide mix, which may be due to differences in epitope activation between both preparations. Surprising responses of some non-infected control subjects to TNF-α stimulation with the wt and alpha mutant JPT mixes may be due to cross-reactivity, as shown in recent studies (29), and will be further investigated in additional studies.

A somewhat superior effect after vaccination with Moderna may be assumed from the data, but did not reach statistical significance in our experimental set-up. It would however corroborate other recently published studies – and may be due to the higher dosing (100 µg) of the Moderna vaccine compared to BioNtech (30 µg)). This interesting topic is being investigated in several other studies (30).

Our results are comparable to other recently published and simultaneous publications (e. g. 15, 31): In our study, measurements over time were not assessed because we could not include longitudinal, repeated measurements of the same subjects. Since broad vaccination had started in March 2021 in Germany due to prior vaccine shortage, and our study was done in July/August 2021, we could only include subjects at an average of 5.2 weeks after the second dosing (minimum: 2 weeks; maximum: 3 months). At the time of the study, no long-term vaccine responses could be obtained. A rapid antibody decline pattern has in the meantime been found by others (e.g. reference 32). This very relevant aspect will be further investigated in future studies.

In contrast, long-term results are included for patients up to 1 year after natural infection. In our set-up, subjects who were investigated 2 months p.i. only showed low anti-RBD binding titers, which does not imply that titers increase over time, since we observed higher titers in the 1 year p.i. group of different individuals. In contrast, other recent pivotal studies showed enhanced neutralizing breadth against SARS-CoV2 1 year after natural infection (15).

The correlation between individual specific CD4+ T cell responses and anti-RBD antibodies was low, probably because B and T cell interaction is not linear, in accordance with previous studies (23). In addition, we also studied the frequencies of CD8+ effector T cells after stimulation with specific peptides. Generally, these frequencies were very low or could not be detected. These results corroborate those of other recent studies on CD4+ and CD8+ T cell responses after stimulation with pools of 15-meric epitope peptides, which we used in our study (12).

Our results demonstrate that a combination of fast expression of recombinant proteins in mammalian cell systems and adaptive T cell assays allows for quick investigation of immune responses to virus variants within 3 months after their emergence. It may be possible within 6 weeks for future mutants. Identification of immune reactions against the current virus mutants and future variants of concern will allow to adapt vaccine strategies in order to provide population protection against the emerging lineages of concern of SARS-CoV-2, and to survey the spread of novel mutants. Our results imply that additional vaccination should be considered after natural infection with SARS-CoV2 in view of global mutant spread. The study summarizes a complementary diagnostic panel to assess immune reactions to vaccines.

## Material and Methods

### Research Subjects

Blood was collected from 67 volunteers with informed consent, who had been previously infected with the SARS-CoV-2 corona virus or been vaccinated against the virus or been unexposed. SARS-CoV-2 infection was confirmed by PCR testing. All vaccinated individuals received two doses of the vaccine and the blood was sampled 5.2±0.9 weeks (minimum: 2 weeks, maximum: 3 months) after the second vaccination (status: completely vaccinated). Mean age of vaccinated subjects was 51±3 years, 10 males and 13 females, and mean age of those who had been naturally infected was 49±2 years, 9 males and 8 females; all were from Caucasian descent. At the time of blood collection, no active infection was present. In the sample we studied, patients had only presented with mild or moderate symptoms, because we did not include patients who had to be hospitalized. This project was submitted for ethical review to and approved by the Bavarian Medical Chamber. Written informed consent was obtained from all subjects, and witnessed by the informing medical doctor. No minors were included.

### SARS-CoV-2-derived Peptides

SARS-CoV-2-derived spike overlapping peptide mixes (15-meric peptides with a 4 amino acid overlap each) from the Wuhan wt strain were purchased from Miltenyi and JPT and from the alpha (B1.1.7), beta (B1.351), gamma (P.1) and delta (B.1.617.2) mutant strains were purchased from JPT (PepMix SARS.SMUT01, SMUT02, SMUT03-1, SMUT06-1). The alpha mutant (B.1.1.7) contains the following mutations: H0069-, V0070-, Y0144-, N0501Y, A0570D, D0614G, P0681H, T0716I, S0982A, D1118H, with the RBD including a single N501Y mutation. The beta (B.1.351) variant spike protein contains the mutations D0080A, D0215G, L0242-, A0243-, L0244-, K0417N, E0484K, N0501Y, D614G, A0701V, of which K417N, E484K and N501Y are located within the RBD. In contrast, the gamma mutant (P.1) is characterized by the following mutations: L018F, T020N, P026S, D138Y, R190S, K417T, E484K, N501Y, D614G, H655Y, T1027I, V1176F, and its RBD displays mutations K417T, E484K and N501Y. The delta variant (B.1.617.2) spike protein is characterized by the following mutations compared to wt: T019R, G142D, E156-, F157-, R158G, L452R, T478K, D614G, P681R and D950N, of which L452R and T478K are part of the RBD. Peptide mixes were dissolved in ddH_2_O with or without 1% DMSO (Roth).

An overview on all peptide mixes is also provided in Table 1.

**Table 1:**
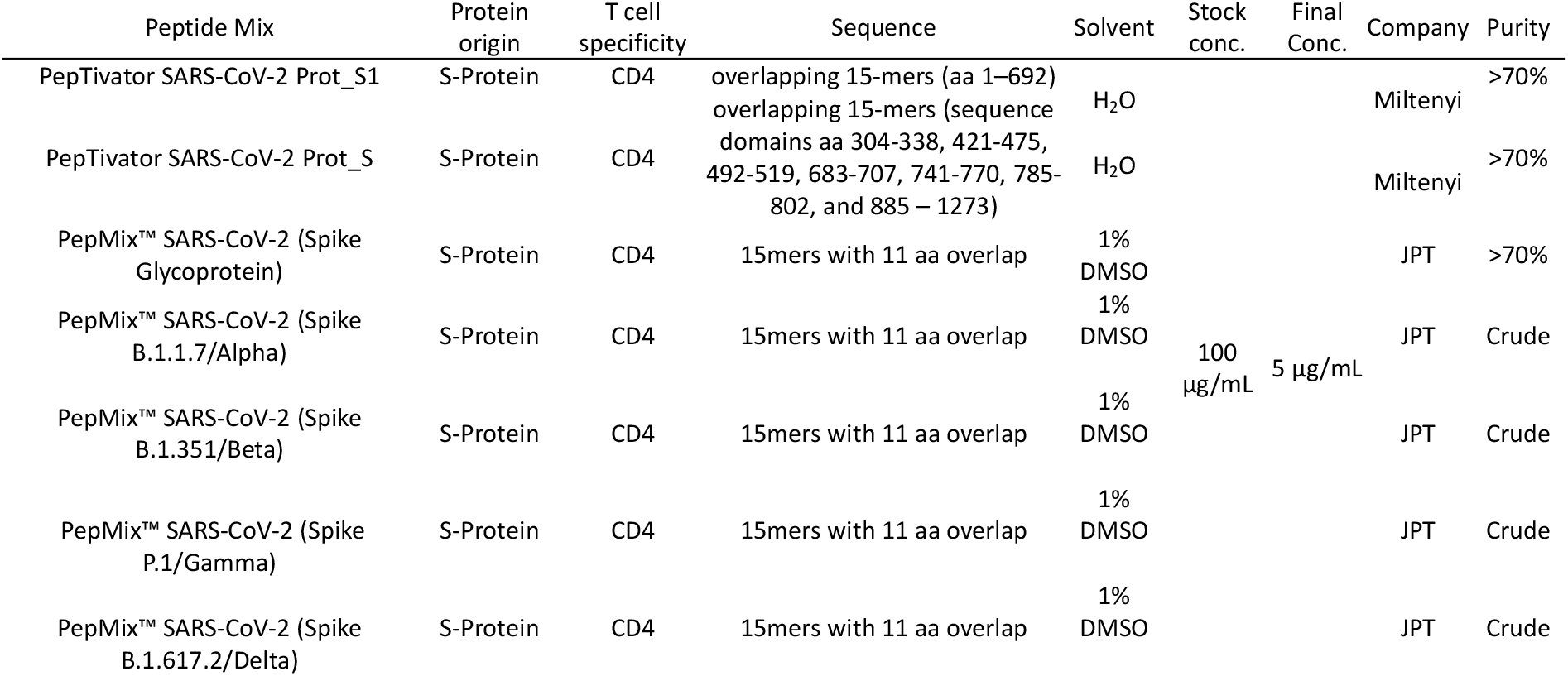
SARS-CoV2 spike protein-derived epitope peptide pools.

### Isolation and Freezing of Peripheral Blood Mononuclear Cells and Serum

Blood was collected for peripheral blood mononuclear cell (PBMC) isolation (in S-Monovettes® K3 EDTA (Sarstedt) or equal), as well as for serum isolation (S-Monovettes® Serum (Sarstedt) or equal). The blood was diluted with an equal volume of PBS (Corning) containing 2% fetal bovine serum (FBS; Gibco; heat-inactivated; from Brazil). PBMCs were obtained by density gradient centrifugation using SepMate™-50 tubes (Stemcell Technologies) filled with Lymphoprep™ density gradient medium (Stemcell Technologies) according to the manufacturer’s instructions. After the isolation, the PBMCs were stained with Acridine Orange and DAPI (solution 18, Chemometec), counted using the NC-250 cell counter (Chemometec), frozen in FBS with 10% DMSO, and stored in liquid nitrogen for at least one week before further experiments. Blood collected for serum isolation was centrifuged 10 min at 2,000xg. Serum was transferred into Eppendorf tubes and frozen at -20°C until use for further experiments.

### T-cell Stimulation and Intracellular Cytokine Staining

*Ex vivo* T-cell responses were analyzed by intracellular cytokine staining (ICS) after stimulation. To this aim, PBMCs were thawed and rested in T cell medium (TCM; RPMI1640 (+GlutaMAX +HEPES, Gibco) + 8% FBS + 1x Penicillin/Streptomycin (Gibco)) containing 1µg/mL DNase (Sigma) for 4-6 hours at 37°C with 5% CO_2_ prior to stimulation. Afterwards, 0.5 - 1.5×10^6^ cells per well (96-well plate) were stimulated with 5 µg/mL of the peptide mixes. DdH_2_O with or without DMSO (depending on the solvent of the peptides) and PMA (10µg/mL)/ Ionomycin (1µM) (both from Sigma) were added as negative and positive controls, respectively. The protein transport inhibitor Brefeldin A (10µg/mL, eBioscience) was added simultaneously and cells were incubated for 14-16h at 37°C, 5% CO_2_. The next day, cells were washed in PBS + 0.5% BSA (Sigma) and stained extracellularly for 20 min at 4°C (Table 1). Thereafter, cells were fixed with IC Fixation Buffer (Intracellular Fixation & Permeabilization Buffer Set, eBioscience) for 30 min at room temperature. Cells were washed and 1x permeabilization buffer (dilute 10x permeabilization buffer from the intracellular fixation & permeabilization buffer set, eBioscience with 9x ddH20) was added and incubated for 5 min, before intracellular staining for 30 min at room temperature (Table 1). Subsequently, cells were washed twice, resuspended in PBS + 0.5% BSA and analyzed with the MACSQuant10 flow cytometer (Miltenyi). At least 200,000 events were counted for each individual condition. The antibodies including clone # which were used for staining were all commercially available, and are listed in Table 2.

**Table 2:** Antibody clones used for FACS analysis.

|  | Marker | Fluorochrome | Clone | Manufacturer | Cat. No. |
| --- | --- | --- | --- | --- | --- |
| Extracellular Staining | Live/dead | fixable Aqua | - | ThermoFisher | L34965 |
|  | CD3 | VioBlue | BW264/56 | Miltenyi | 170-081-046 |
|  | CD4 | APC | VIT4 | Miltenyi | 130-113-210 |
|  | CD8 | PerCP | BW135/80 | Miltenyi | 130-113-160 |
| Intracellular Staining | IFN | PE | 45-15 | Miltenyi | 130-113-493 |
|  | CD154 | FITC | 5C8 | Miltenyi | 130-113-606 |
|  | TNF | PE-Vio770 | cA2 | Miltenyi | 130-120-492 |
|  | IL-2 | APC-Vio 770 | REA689 | Miltenyi | 130-111-492 |

Representative FACS images are shown in Figure 8, which also illustrates the gating strategies.

**Figure 8:**
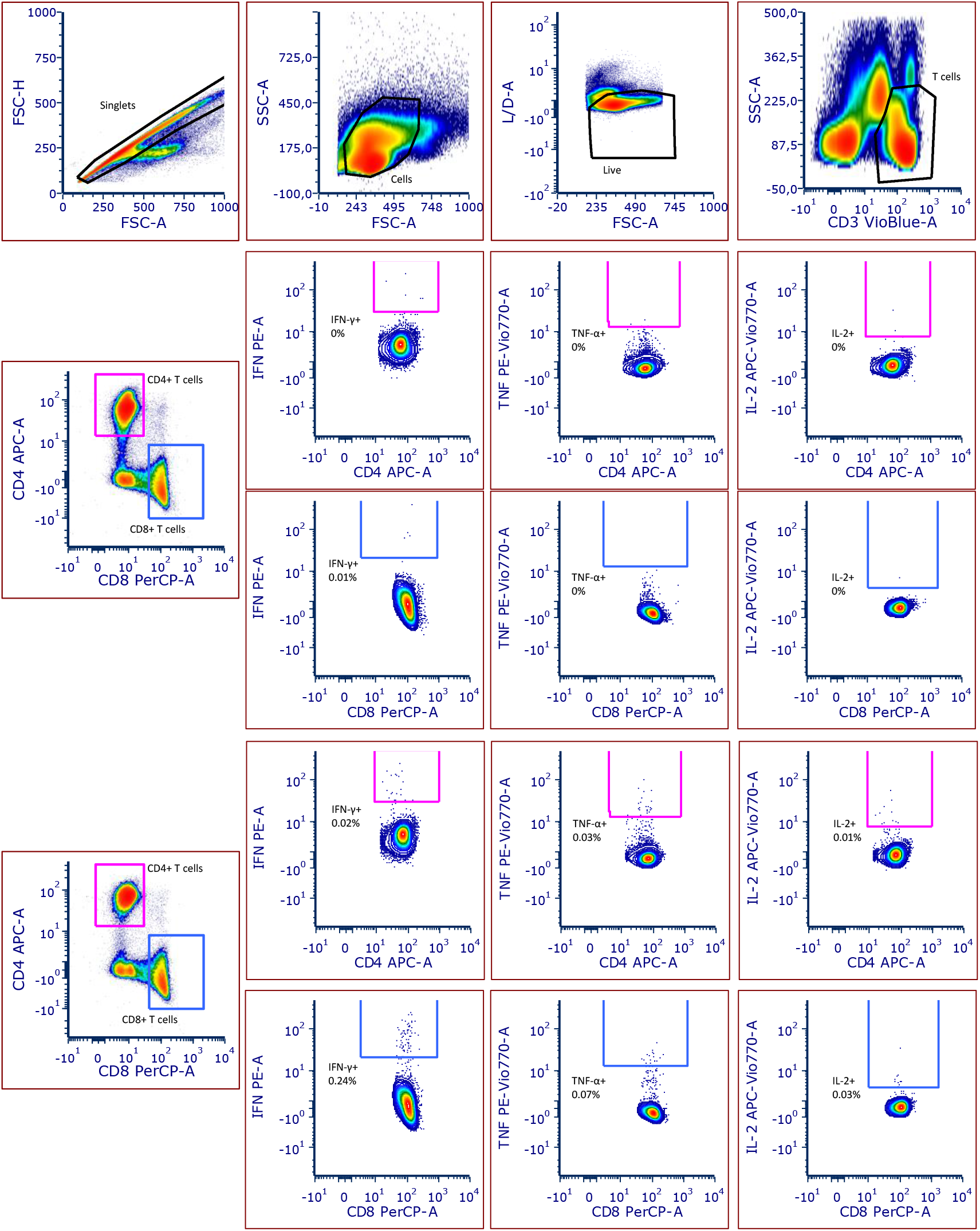
Gating strategy of the intracellular cytokine staining. The figure shows a representative flow cytometry gating strategy for one of the donors. Cells were gated as follows: Singlets, Cells, Live, CD3+ T cells (A) and further T cells were divided into CD4+ and CD8+ T cells and from each of the subsets IFN-γ+, TNF-α+, or IL-2+ cells were gated (B, C). Panel B shows the negative control and Panel C shows the restimulated condition with the Miltenyi peptide mix of the same representative donor.

The data analysis was performed with FCS Express 7 (V. 7.06, De Novo Software). All results were audited, and responses were defined as positive if the percentage of marker-positive cells after peptide stimulation was ≥two-fold compared to the negative control and >20 marker-positive cells were recorded. The frequency of the negative controls was subtracted from the frequency of marker-positive cells.

### Generation of CHO RBD-His and CHO-NP-Strep Cell Lines

S protein RBD-His consists of the amino acids corresponding to the receptor binding (RBD) domain, which was derived from the S protein nucleotide sequence (positions 22517-23183, amino acids 319 to 541, RVQP….CVNF) of the SARS-CoV2 Wuhan Hu-1 genome (Genbank accession number MN908947) followed by six histidines. Nucleocapsid protein N-strep consists of the amino acids corresponding to the N protein nucleotide sequence (positions 28290 to 29549) of the SARS-CoV2 Wuhan Hu-1 genome (Genbank accession number MN908947) followed by a streptavidin tag (NP-Strep). Accordingly, alpha (B.1.1.7) RBD contains a single N501Y mutation, whereas the beta (B.1.351) variant RBD displays K417N, E484K and N501Y and gamma (P.1) RBD mutations K417T, E484K and N501Y. In contrast, the delta variant (B.1.617.2) spike RBD contains L452R, T478K.

The complementary DNA sequences adapted for hamster codon usage were produced synthetically by GeneArt (Life Technologies) by adding the signal sequence METPAQLLFLLLLWLPDTTG at the beginning and cloned into the plasmid vector pcDNA5/FRT via BamHI and XhoI. The resulting vectors were called pcDNA5/CoV-RBD-His and pcDNA5/CoV-NP-Strep, respectively, and allow for expression and secretion of RBD-His or NP-Strep into the culture medium of mammalian cells under the control of the human cytomegalovirus (CMV) immediate-early enhancer/promoter. The selection for stable clones was done using Hygromycin B after co-transfection with the plasmid pOG44. The vectors were transfected into Flip-InTM-Chinese hamster ovary (CHO) cells (Life Technologies) using the Lipofectamine 2000 Reagent (Invitrogen, #11668-019), together with the plasmid pOG44, providing site-directed recombination. After selection of a stably expressing clone for each line in Ham’s F12 medium supplemented with 10% FBS and 600 µg/ml Hygromycin B, the clones were adapted to ProCHO5 medium (Lonza, #BE12-766Q) supplemented with 4 mM L-glutamine (Biochrom, #K0283).

### Expression and purification of RBD-His and NP-Strep

The various CHO-spike-RBD-His cells and CHO-spike-NP-Strep cells were grown in suspension in ProCHO5, 4 mM L-glutamine and 600 µg/ml Hygromycin B in flasks to submaximal density at 37°C. The supernatants were collected after centrifugation at 400xg for 5 min and subsequent filtration with a 0.22 nm sterile filter (TPP, #99722). The resulting RBD-His or NP-Strep protein-containing medium was immediately frozen and stored at -20°C until protein purification. The cells were continuously grown at 37°C, with splitting and supernatant collection every 3 – 4 days.

For protein purification, thawed CHO-RBD-His supernatants (1 L) were loaded onto an equilibrated 1 ml HisTrapTM excel column (GE Healthcare 17-3712-05). After washing the column with wash buffer (20 mM sodium phosphate, 0.3 M NaCl, pH 8.0), RBD-His was eluted with 4 × 1 ml 0.25 M imidazole, 20 mM sodium phosphate, 0.3 M NaCl, pH 8.0. Protein concentration and, hence, content was determined by OD 280 measurement and the relevant fractions were dialyzed (Slyde-A-Lyzer Dialysis Cassette, 10000 MWCO, Thermo Scientific # 66380) against phosphate-buffered saline (PBS from Roth: 137 mM NaCl, 2.7 mM KCl, 10 mM Na2HPO4, 2 mM KH2PO4, pH 7.4, 0.2 μm filtered and steam sterilized) at 4°C for 16 h. 0.5 L CHO-NP-Strep supernatants were diluted 1:2 in 50 mM sodium phosphate, 0.3 M NaCl, pH 8.0, and loaded on an equilibrated 1 ml StrepTrapTM HP column (GE Healthcare 28-9075-46). After washing the column with 50 mM sodium phosphate, 0.3 M NaCl, pH 8.0, NP-Strep was eluted with 4 × 1 ml 20 mM sodium phosphate, 0.3 M NaCl, 2.5 mM desthiobiotin (Sigma, # D 1411) pH 8.0. Protein content was determined by OD 280 measurement and the relevant fractions were dialyzed (Slyde-A-Lyzer Dialysis Cassette, 10000 MWCO, Thermo Scientific # 66380) against PBS at 4°C for 16 h.

For biotinylation of RBD, delta mutantRBD or ACE2, the EZ-Link Micro Sulfo-NHS-LC biotinylation kit (Thermo Scientific # 21935 or #A39257) was used with 20-fold molar excess according the standard procedure instruction, followed by removal of excess biotin by dialysis against 1 l PBS for 16 h at 4°C, using Slide-A-Lyzer™ Dialysis Cassettes, 7K MWCO, 0.5 mL (ThermoFisher # 66373).

### Qualitative Detection of anti-SARS-CoV2 Antibodies in Human Serum

Anti-SARS-CoV2 antibodies in human serum or in a drop of full blood were detected by a two-step incubation antigen sandwich ELISA using the RBD of the S1 protein of the SARS-CoV2 virus. All procedures were performed at room temperature (RT) and incubations were done on a microtiter plate shaker. ELISA plates were coated with 60 µl/ well RBD-His protein (final concentration 0,45 µg/ml) in coating solution (Candor, #121125) for 1 h. The coated plates were washed three times with PBST (PBS, 0.1 % Tween-20), blocked with 100 µl/well of blocking solution (PBST, 3 % milk powder) for 1 h, and washed again. 30µl serum samples were diluted 1:2, 1:20, 1:200 and 1:2000 with PBS and transferred to the blocked ELISA plates and incubated for 1h. After washing three times with PBST, the ELISA plates were incubated for 1h with 60 µl/well of biotinylated RBD-His (Bio-RBD-His, final concentration 0.02 µg/ml, diluted in PBST). The plates were washed three times with PBST and incubated with Strep-POD (Jackson Immunoresearch, #016-030-084) diluted in PBST 1:50000 for 1h. After washing five times, bound POD was detected by incubation with 100 µl/ well of TMB substrate (Thermo Scientific, #34029) until a maximal optical density (OD) of about 1 to 2 was reached. Finally, the colorimetric reaction was stopped with 100 µl/ well stopping solution (1M H_2_SO_4_) and the OD determined at a wavelength of 450 nm with a reference wavelength of 595 nm in a plate reader.

### Neutralisation of wt RBD-Biotin binding vs. delta mutant RBD-Biotin binding to immobilized ACE2-His by serum antibodies

All procedures were performed at room temperature (RT) and incubations were done on a microtiter plate shaker. ELISA plates were coated with 60 µl/ well ACE2-His protein with 1 µg/ml in PBS for 1 h. The coated plates were washed three times with PBST (PBS, 0.1 % Tween-20), blocked with 100 µl/well of blocking solution (PBST, 3 % milk powder) for 1 h, and washed again. 60µl of RBDwt-Bio (0,3µg/ml) or RBDdelta-Bio (0,3µg/ml) were co-incubated with various serum sample dilutions (1:2, 1:4, 1:8, 1:16, 1:48, 1:160, 1:1480, 1:1600 in PBST) and transferred to the blocked ELISA plates and incubated for 1h. The plates were washed three times with PBST and incubated with Strep-POD (Jackson Immunoresearch, #016-030-084) diluted in PBST 1:20000 for 1h. After washing four times, bound POD was detected by incubation with 100 µl/ well of TMB substrate (Thermo Scientific, #34029) until a maximal optical density (OD) of about 1 to 2 was reached. Finally, the colorimetric reaction was stopped with 100 µl/ well stopping solution (1M H2SO4) and the OD determined at a wavelength of 450 nm with a reference wavelength of 595 nm with in a plate reader.

Then, IC50 values minus blank values were calculated for each sample using results from all dilutions with SigmaPlot v14.5. These IC50 values represent neutralizing serum dilutions. Ratios between IC50s calculated for inhibition of wt vs. delta mutant RBD binding to immobilized ACE2 were used for further analysis.

### Neutralisation of ACE2-Biotin binding to immobilized wt RBD vs. delta mutant RBD by serum antibodies

All procedures were performed at room temperature (RT) and incubations were done on a microtiter plate shaker. ELISA plates were coated with 60 µl/ well RBDwt protein or RBDdelta with 2 µg/ml in coating buffer (50mM carbonate buffer, Na2CO3/NaHCO3, pH9.6) for 1 h. The coated plates were washed three times with PBST (PBS, 0.1 % Tween-20), blocked with 100 µl/well of blocking solution (PBST, 3 % milk powder) for 1 h, and washed again. 60µl of ACE2-Bio with 0,3µg/ml was co-incubated with various serum sample dilutions (1:2, 1:4, 1:8, 1:16, 1:48, 1:160, 1:1480, 1:1600 in PBST) and transferred to the blocked ELISA plates and incubated for 1h. The plates were washed three times with PBST and incubated with Strep-POD (Jackson Immunoresearch, #016-030-084) diluted in PBST 1:20000 for 1h. After washing four times, bound POD was detected by incubation with 100 µl/ well of TMB substrate (Thermo Scientific, #34029) until a maximal optical density (OD) of about 1 to 2 was reached. Finally, the colorimetric reaction was stopped with 100 µl/ well stopping solution (1M H2SO4) and the OD determined at a wavelength of 450 nm with a reference wavelength of 595 nm with in a plate reader.

Then, IC50 values minus blank values were calculated for each sample using results from all dilutions with SigmaPlot v14.5. These IC50 values represent neutralizing serum dilutions. Ratios between IC50s calculated for inhibition of ACE2 binding to immobilized wt vs. delta mutant RBD were used for further analysis.

### Neutralisation of ACE2-Biotin binding to immobilized omicron mutant RBD vs. delta mutant RBD by serum antibodies

All procedures were performed at room temperature (RT) and incubations were done on a microtiter plate shaker. ELISA plates were coated with 60 µl/well RBDomicron protein or RBDdelta with 1 µg/ml in coating buffer (50mM carbonate buffer, Na2CO3/NaHCO3, pH9.6) for 1 h. The coated plates were washed three times with PBST (PBS, 0.1 % Tween-20), blocked with 100 µl/well of blocking solution (PBST, 3 % milk powder) for 1 h, and washed again. 60µl of ACE2-Bio with 0,3µg/ml was co-incubated with various serum sample dilutions (1:2, 1:6, 1:20, 1:60, 1:200, 1:600, 1:2000, 1:20000 in 2x PBST) and transferred to the blocked ELISA plates and incubated for 1h. The plates were washed three times with PBST and incubated with Strep-POD (Jackson Immunoresearch, #016-030-084) diluted in PBST 1:20000 for 1h, and further processed as outlined above.

### Statistical Analysis

Statistical analysis was performed using SPSS software version 26. Statistical differences were determined using one-way ANOVA with Tukey’s post-hoc test in case data were normally distributed (as assessed by Shapiro-Wilk normality test). Otherwise, data were analyzed using the Kruskal-Wallis nonparametric test.

## Data Availability

All data are available with the authors

## Author’s Contributions

JRR, HPH conceived the studies, carried out experiments, analysed data and wrote parts of the manuscript. RG and VM carried out experiments and analysed data. MU and ML conceived the parts of the studies and wrote and revised the manuscript and revisions.

## Acknowledgments

Kerstin Dehne and Markus Kraller

